# Neonatal and Maternal Outcome of COVID-19 positive women in Sri Lanka: Secondary Analysis using National COVID-19 Positive Pregnant Women Surveillance

**DOI:** 10.1101/2022.02.10.22270732

**Authors:** Malith Kumarasinghe, Kaushalya Kasturiaratchi, Hemali Jayakody, Shakira Irfaan, Wasana Samarasinghe, Harendra Dassanayake, Sanjeeva Godakandage, Chithramalee de Silva

**Author notes:** No 54/A, New Jayaweera Road, Ethul-Kotte, Sri Jayawardhanapura Kotte 10100.

## Abstract

**Objectives:** This study aims to describe the population level data on neonatal and maternal outcomes of COVID-19 positive pregnant women of Sri Lanka by secondary analysis using National COVID-19 Positive Pregnant Women Surveillance.

**Design:** Secondary analysis of surveillance data from the National COVID-19 positive pregnant women surveillance, Sri Lanka. Data of all pregnant women whose maternal and neonatal outcomes were reported in National Surveillance from 1st March 2020 to 31st October 2021 were included in the study. Associated factors for maternal and neonatal outcomes, namely POA at delivery, mode of delivery, birthweight, immediate place of newborn care, congenital abnormalities, and condition of neonate at completion of one month were calculated using univariate and multivariate Odds ratios.

**Results:** Maternal COVID-19 infection reported preterm birth rate of 11.9%, LSCS rate of 54.5%, low birthweight rate16.5% and 8.3% of the newborns requiring intensive care. Neonatal mortality rate was 9 per 1000 live births. Pre-pregnancy overweight and obesity increased the risk of preterm delivery compared to pregnant women with normal BMI by 46.7% (AOR=1.467, CI=1.111-1.938, P=0.007). In contrast, the risk of preterm delivery reduced by 82.4% (AOR=0.176, CI=0.097-0.317, p<0.001) and presence of any type of congenital abnormalities in newborns by 72.4% among the COVID-19 positive women who required only inward treatment in comparison to women with severe COVID-19 infection requiring intensive care (AOR=0.276, CI=0.112-0.683, p=0.005).

**Conclusion:** Increased severity of maternal COVID-19 infection and pre-pregnancy overweight/ obesity were associated with many adverse pregnancy and neonatal outcomes. Therefore, close observation and aggressive management of COVID-19 among the pregnant women should be considered to reduce the risk of progressing to severe illness.

## INTRODUCTION

In 2019, the novel coronavirus SARS-CoV-2 pandemic originated in Hubei Province of People’s Republic of China. Within few months COVID-19 spread globally with majority of nations reporting cases. WHO Emergency Committee declared COVID-19 as a global health emergency on 30^th^ January 2020 and 11th March 2020 as a pandemic.[1]

At the early stages of the pandemic, minimal knowledge existed on pregnancy and newborn outcomes of maternal COVID-19 infection unlike the understanding on disease severity and clinical profile. However, with the continuation of the pandemic, evidence on maternal and newborn outcomes in maternal COVID-19 gradually emerged. Many studies have highlighted the possibility of numerous adverse maternal and perinatal outcomes following COVID-19 infection.[2-6] Possible adverse maternal and perinatal outcomes include increased risk of preeclampsia, maternal death, disseminated intravascular coagulopathy, intrauterine fetal demise, intrauterine growth restriction, preterm delivery, spontaneous abortion, preterm birth and stillbirth. Further, systematic reviews further concluded that severe maternal COVID-19 infection significantly increase these adverse outcomes compared to mild to moderate infection.

An overview of systematic reviews reported that cesarean section is the preferred mode of delivery which ranged from 52.3% to 95.8%.[7] Further, the study team reported the preterm delivery rate between 14.3% to 63.8%. Stillbirth rate reported to be less than 2.5% and low birthweight between 5.3% to 47.4%. NICU admission of the newborns of COVID-19 positive pregnant women ranged from 3.1% to 76.9% with neonatal mortality rate did not exceed 3%. Despite the inclusion of 39 systematic reviews, the wide range of rates in pregnancy and neonatal outcomes highlight the non-agreement of the scientific community on the effect of maternal COVID-19 on pregnancy and neonatal outcomes.[7]

The literature revealed wide range in rates of pregnancy and neonatal outcomes in maternal COVID-19 infection, not only among the different studies but also in different geographical locations.[8-9] Systemic review by Dubey et al. revealed that deliveries by cesarean sections were higher among Chinese communities in comparison to American and European communities (91% vs. 40% and 38% respectively). Similarly higher rates of adverse pregnancy outcomes were reported in Chinese studies compared to American and European studies (21% vs 15% and 19% respectively). Further, the same systematic review showed reginal variation in preterm delivery. (American studies vs. Chinese and European studies: 12% vs. 17% and 19% respectively).[8] These findings highlight the importance of local evidence for the management of pregnancy and neonatal outcomes of maternal COVID-19.

Though scientific information has emerged in developed countries on impact on COVID-19 maternal infection on pregnancy and newborn outcomes, the scarcity of information in developing countries, particularly in South Asia is a concern. In Sri Lanka, no literature is available on the pregnancy and newborn outcomes following maternal COVID-19 infection. Further, limited understanding prevails on factors which increase the risk of adverse pregnancy and neonatal outcomes in maternal COVID-19 infection. Therefore, this study was intended to answer 3 main questions related to pregnancy and newborn outcomes following maternal COVID-19 in Sri Lanka. Number one: To describe the basic sociodemographic, maternal, pregnancy and COVID-19 disease related characteristics of COVID-19 positive pregnant women in Sri Lanka. Number two: To describe the factors significantly associated with the selected pregnancy outcomes among the COVID-19 positive pregnant women in Sri Lanka. Number three: To describe the factors significantly associated with the neonatal outcomes of COVID-19 positive pregnant women in Sri Lanka.

In this study, we aimed to describe the maternal, neonatal and COVID-19 disease related characteristics among the COVID-19 positive pregnant women in Sri Lanka using data from national COVID-19 positive pregnant women surveillance. In addition, the association of pregnancy and neonatal outcome variables were assessed with selected sociodemographic, maternal, pregnancy and COVID-19 disease related factors.

## METHODS

Secondary analysis was conducted using the data available at National COVID-19 positive pregnant women surveillance in Sri Lanka.

### National COVID-19 positive pregnant women surveillance

National COVID-19 positive pregnant women surveillance was initiated by the Ministry of Health to collect the information of COVID-19 positive pregnant women. The directive issued by the Director General of Health Services of Sri Lanka on 07th June 2021 instructed all health staff to register and follow up all COVID-19 positive pregnant women in Sri Lanka from 2020 onwards.[10] Therefore, data on positive women from the onset of the first case in Sri Lanka was gathered retrospectively and it is expected to continue until 2022. Information on COVID-19 positive pregnant women were reported by over 6000 field Public Health Midwife (PHM) in the country using a data extraction form. End of each month, the data was transferred to the electronic Reproductive Health Management Information System (eRHMIS) of Family Health Bureau. eRHMIS is the current electronic data management system of Reproductive, Maternal, New-born, Child, Adolescent, and Youth Health (RMNCAYH) programme in Sri Lanka based on the DHIS2 platform. The database included data of pregnant women who were tested positive either by RT-PCR or by Rapid Antigen Test for SARS-CoV-2 virus.

Data extraction form comprised of four segments. First segment included socio-economic data, pregnancy details and other co-morbidities of all COVID-19 positive pregnant women. Segment two comprised of follow up information at the time of discharge. This included details on the severity of COVID-19 infection in women during the hospital stay and the pregnancy status at the time of discharge. PHM followed up the pregnant woman in the field and completed the third segment of the form at one month postpartum. It comprised information on both maternal and neonatal outcome. Fourth segment contained information of postnatal assessment of the infant at 6 months. The results of the 6 months assessment will be analysed and disseminated in future.

### Study variables

#### Socio-demographic characteristics

Age in years (grouped according to less than 35 years and 35 years or more), ethnicity, sector (urban, rural and estate according to the definition by the department of Census and Statistics) and monthly family (household) income according to the poverty line, median and mean household income.[11-12].

#### Maternal characteristics

Selected maternal characteristics include the pre-pregnancy BMI grouped as low, normal and overweight (and obesity), comorbidities, namely chronic DM, chronic hypertension, cardiovascular disease, chronic respiratory disease.

#### Pregnancy and neonatal characteristics

Pregnancy characteristics selected for the study include the presence of gestational DM, pregnancy induced HT, gravida, parity, number of living children, POA at delivery (preterm birth or not), mode of delivery (vaginal delivery or cesarian section). Neonatal characteristics include birthweight grouped according to WHO classification, place of immediate newborn care (postnatal ward or premature baby unit/ neonatal intensive care unit), initiation of breast feeding within one hour of birth, detection of any congenital abnormalities and neonate live or not at the end of neonatal period.[13]

#### COVID-19 related measures

COVID-19 vaccination status was categorized according to non-vaccinated or received at least single dose of any COVID-19 vaccination before 14 days of the diagnosis of COVID-19. Time of diagnosis of COVID-19 during the pregnancy was classified according to the trimester. Severity of the COVID-19 infection was categorized according to the place of the management (all COVID-19 positive pregnant women mandate inward care as per Department of Health of Sri Lanka).[14]

### Study methods and analysis

Data extraction of all eligible COVID-19 positive pregnant women was carried out from the national surveillance (Eligibility-In the national surveillance, completed segments number one, two and three of the data extraction form by 31st October 2021) was extracted. Data was initially transferred to MS Excel software; they were coded and analysed using Statistical Package for Social Sciences (SPSS) 22 version. Rates were presented as proportions. Statistical significance was calculated based on a p value of less than 0.05 was considered as statistically significant. All variables were used to perform the multivariate analysis using logistic regression (enter method) irrespective of their significance in bivariate analysis to identify multivariate ORs and their Confidence Intervals. For monthly family (household) income, poverty line, median and mean household incomes were used for categorization.

## RESULTS

### Description of the of the study population

Total of 9905 pregnant women were identified as COVID-19 positive in National COVID-19 Positive Pregnant Women Surveillance by 31^st^ of October 2021. Majority of these cases were diagnosed in the 3^rd^ quarter of 2021 and therefore the delivery outcome of most of these women was not occurred at the time of the analysis. Thus, only in 2493 COVID-19 positive pregnant women, details of pregnancy and neonatal outcomes were available by 31st October 2021 in the surveillance. Out of these 2493 cases, 153 pregnant women reported miscarriages (1^st^ and 2^nd^ trimester) and was excluded from the analysis. Therefore, pregnancy and neonatal outcomes of 2340 COVID-19 positive pregnant women were included for the analysis.

Close to quarter of the COVID-19 positive pregnant women were below the poverty line (24.1%).[12] Further, 80% of the pregnant women were below the median monthly household income of Sri Lanka.[12] Close to 18% of the pregnant women were 35 years or more (17.9%). Pre-pregnancy overweight among the COVID-19 positive pregnant women stood at 35.7%. Only 31% reported receiving at least single dose of any type of COVID-19 vaccine. Incidence of Gestational DM was at 8% whereas Pregnancy Induced Hypertension was reported as 3% among the COVID-19 positive pregnant women (Table 1).

**Table 1.**
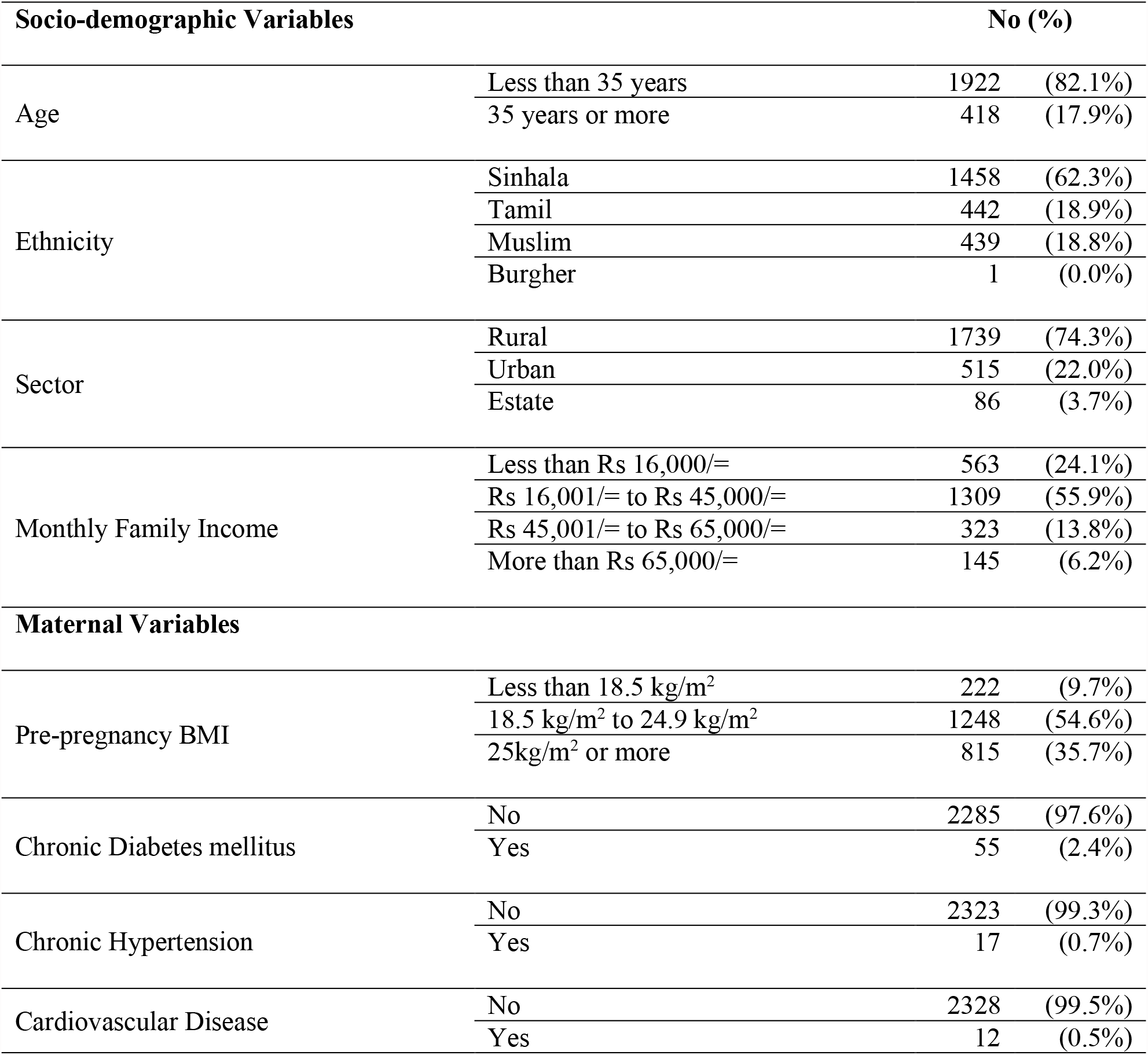
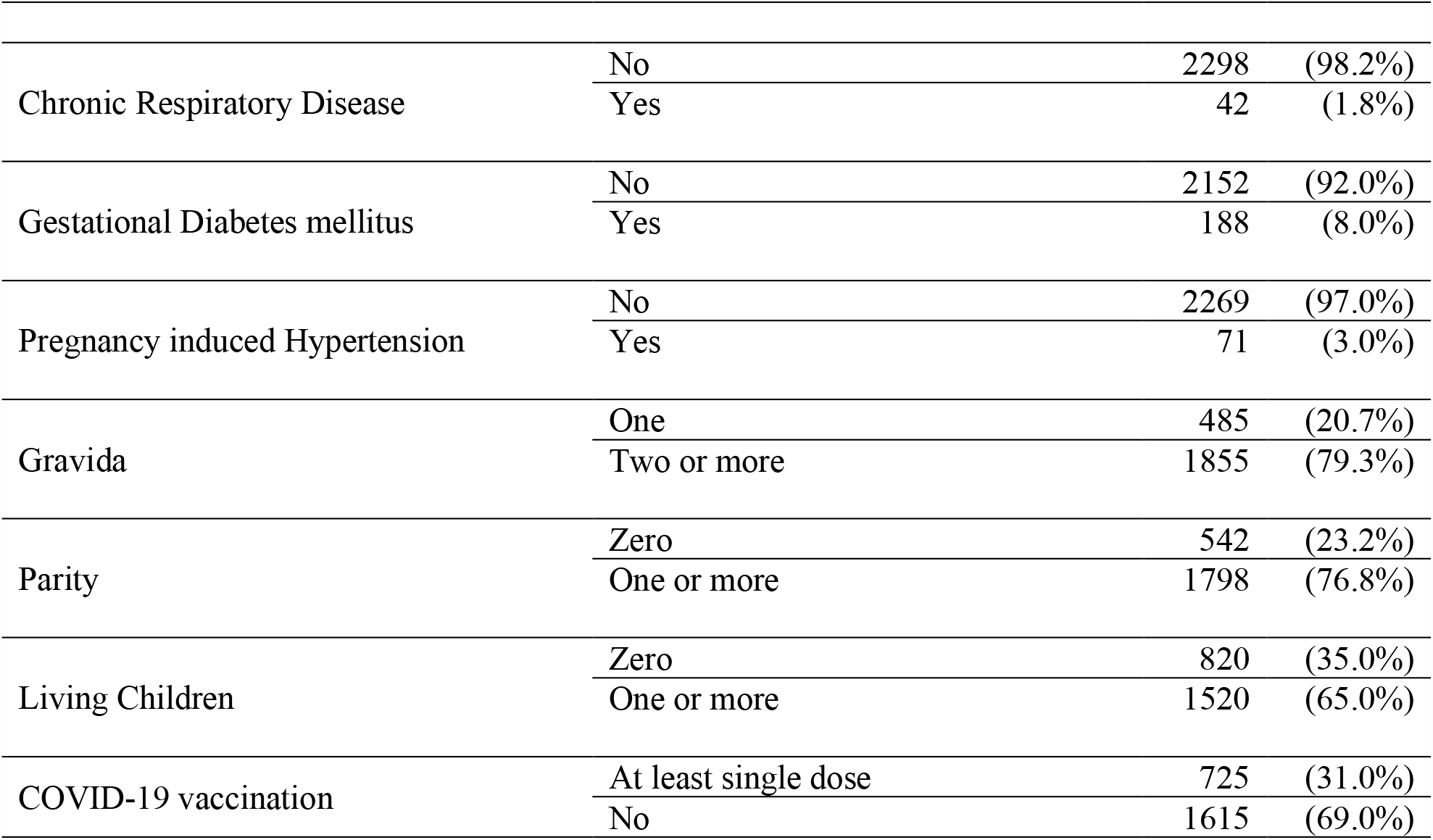
Socio-demographic and maternal characteristics of COVID-19 positive pregnant women of Sri Lanka.

COVID-19 positive pregnant women showed preterm birth of 11.9% and LSCS rate at 54.5%. Low birthweight rate was 16.5% with 8.3% of the newborns were admitted to either Premature Baby Unit or Neonatal Intensive Care Unit. Neonatal mortality rate was reported at 9 per 1000 live births among the COVID-19 positive pregnant women (Table 2).

**Table 2.** Pregnancy and neonatal characteristics of COVID-19 positive pregnant women of Sri Lanka.

| Pregnancy Variables |  | No (%) |  |
| --- | --- | --- | --- |
| POA at Delivery | 37 weeks or more | 2043 | (88.1%) |
|  | Less than 37 weeks | 276 | (11.9%) |
| Mode of Delivery | Normal Vaginal Delivery | 1028 | (44.3%) |
|  | Assisted Vaginal Delivery | 26 | (1.1%) |
|  | Elective LSCS | 838 | (36.1%) |
|  | Emergency LSCS | 427 | (18.4%) |
| Neonatal Variables |  |  |  |
| Birth Weight | Extremely Low Birthweight | 8 | (0.3%) |
|  | Very Low Birthweight | 12 | (0.5%) |
|  | Low Birthweight | 363 | (15.7%) |
|  | Normal Birthweight | 1934 | (83.5%) |
| Place of Newborn Care following Birth | Postnatal Ward only | 2135 | (91.7%) |
|  | Premature Baby Unit/ Neonatal Intensive Care Unit | 194 | (8.3%) |
| Breast Feeding Initiated within One Hour of Birth | Yes | 2267 | (97.3%) |
|  | No | 62 | (2.7%) |
| Congenital Abnormalities detected within Neonatal Period | No | 2265 | (96.8%) |
|  | Yes | 75 | (3.2%) |
| Live Neonatal at the end of 1<sup>st</sup> 28 days following birth | Yes | 2319 | (99.1%) |
|  | No | 21 | (0.9%) |

For OR calculations in Table 3 to 7, above median to mean household income (Rs 45,001.00 to Rs 65,000.00) and above the mean household income groups were combined (More than 65,000.00). There was only single Burger COVID-19 positive pregnant woman. The said woman’s mother tongue was Sinhala. Therefore, for OR calculation, the Burger pregnant woman was included to Sinhala ethnicity.(Burger population percentage is only 0.2% of Sri Lankan population according to census 2012).[11]

**Table 3.** Odds Ratios for factors associated with POA at delivery of COVID-19 positive pregnant women. Univariate and multivariate analysis.

|  |  |  |  |  |  |
| --- | --- | --- | --- | --- | --- |
| **Outcome variable- POA at Delivery** |  | **37 weeks or more** | **Less than 37 weeks** | **OR (Univariate)** | **OR (Multivariate)** |
| **0 - 37 weeks or more** |  |  |  |  |  |
| **1 – Less than 37 weeks** |  |  |  |  |  |
| Age | 35 years or more | 350(84.7%) | 63(15.3%) |  |  |
|  | Less than 35 years | 1693(88.8%) | 213(11.2%) | 0.699(0.516-0.947, p=0.021) | 0.786(0.565-1.095, p=0.154) |
| Ethnicity | Tamil | 382(87.6%) | 54(12.4%) |  |  |
|  | Muslim | 383(88.2%) | 51(11.8%) | 0.942(0.626-1.417, p=0.774) | 0.898(0.578-1.394, p=0.631) |
|  | Sinhala* | 1278(88.2%) | 171(11.8%) | 0.947(0.683-1.312, p=0.742) | 0.852(0.589-1.231, p=0.393) |
| Sector | Urban | 448(87.8%) | 62(12.2%) |  |  |
|  | Estate | 78(90.7%) | 8(9.3%) | 0.741(0.342-1.608, p=0.448) | 0.733(0.318-1.691, p=0.467) |
|  | Rural | 1517(88.0%) | 206(12.0%) | 0.981(0.725-1.328, p=0.902) | 1.009(0.732-1.391, p=0.956) |
| Monthly Family Income | Rs 45,001/= or more | 398(85.8%) | 66(14.2%) |  |  |
|  | Less than Rs 16,000/= | 484(86.7%) | 74(13.3%) | 0.922(0.645-1.318, p=0.656) | 0.960(0.645-1.431, p=0.843) |
|  | Rs 16,000/= to Rs 45,000/= | 1161(89.5%) | 136(10.5%) | 0.706(0.515-0.968, p=0.031) | 0.744(0.533-1.038, p=0.082) |
| Pre-Pregnancy BMI | 18.5 kg/m<sup>2</sup> to 24.9kg/m<sup>2</sup> | 1112(89.7%) | 127(10.3%) |  |  |
|  | Less than 18.5 kg/m<sup>2</sup> | 202(91.4%) | 19(8.6%) | 0.824(0.497-1.364, p=0.451) | 0.845(0.505-1.412, p=0.519) |
|  | 25kg/m<sup>2</sup> or more | 684(84.9%) | 122(15.1%) | 1.562(1.197-2.038, p=0.001) | 1.467(1.111-1.938, p=0.007) |
| Presence of Co-morbidities | No | 1703(88.6%) | 220(11.4%) |  |  |
|  | Yes | 340(85.9%) | 56(14.1%) | 1.275(0.930-1.748, p=0.131) | 1.084(0.775-1.515, p=0.637) |
| Gravida | One | 436(90.3%) | 47(9.7%) |  |  |
|  | Two or more | 1607(87.5%) | 229(12.5%) | 1.322(0.949-1.841, p=0.099) | 1.176(0.497-2.783, p=0.712) |
| Parity | One or more | 1558(87.5%) | 222(12.5%) |  |  |
|  | Zero | 485(90.0%) | 54(10.0%) | 0.781(0.571-1.070, p=0.124) | 1.012(0.417-2.456, p=0.979) |
| Living Children | One or more | 1318(87.5%) | 189(12.5%) |  |  |
|  | Zero | 725(89.3%) | 87(10.7%) | 0.837(0.639-1.096, p=0.195) | 1.074(0.707-1.630, p=0.739) |
| POA at Diagnosis | 3<sup>rd</sup> Trimester | 1738(88.2%) | 233(11.8%) |  |  |
|  | 1<sup>st</sup> Trimester | 32(80.0%) | 8(20.0%) | 1.865(0.849-4.095, p=0.121) | 2.069(0.920-4.655, p=0.079) |
|  | 2<sup>nd</sup> Trimester | 273(88.6%) | 35(11.4%) | 0.956(0.656-1.395, p=0.817) | 1.086(0.727-1.622, p=0.688) |
| COVID-19 Vaccination Status | No | 1423(88.8%) | 179(11.2%) |  |  |
|  | At least Single Dose | 620(86.5%) | 97(13.5%) | 1.244(0.955-1.620, p=0.106) | 1.244(0.937-1.652, p=0.132) |
| Severity of COVID-19 Infection | ICU/ECMO/Ventilated/Death<sup>a</sup> | 32(59.3%) | 22(40.7%) |  |  |
|  | Inward care only | 2011(88.8%) | 254(11.2%) | 0.184(0.105-0.321, p<0.001) | 0.176(0.097-0.317, p<0.001) |

\*Includes a single participant of Burger ethnicity (mother tongue was Sinhala)
<sup>a</sup>ICU- intensive care unit; ECMO- extracorporeal membrane oxygenation

### Factors associated with preterm delivery of COVID-19 positive pregnant women

Table three describes the selected sociodemographic, COVID-19, maternal, pregnancy and neonatal variables influence the time of delivery among COVID-19 positive pregnant women of Sri Lanka. Pre-pregnancy overweight and obesity increased the risk of preterm delivery among the COVID-19 positive women compared to normal BMI pregnant women by 46.7% (AOR=1.467, CI=1.111-1.938, P=0.007). In contrast the risk of preterm delivery reduced by 82.4% among the COVID-19 positive women who required only inward treatment compared to the women needing ICU care or died due to COVID-19 (AOR=0.176, CI=0.097-0.317, p<0.001).

### Factors associated with delivery by cesarean section of COVID-19 positive pregnant women

Interestingly, lower income COVID-19 positive pregnant women (equal or less than monthly median family income) were less likely to undergo cesarean section (than vaginal delivery) compared to high income pregnant women (more than monthly median family income) (AOR=0.546, CI=0.413-0.722, p<0.001 and AOR=0.578, CI=0.459-0.729, p<0.001). In addition, overweight or obese COVID-19 positive pregnant women demonstrated higher chance of undergoing cesarean section compared to normal BMI women with 23.3% increase (AOR=1.233, CI=1.021-1.490, p=0.030). In contrast, COVID-19 positive pregnant women with low pre-pregnancy BMI decreased the chances of undergoing cesarean section in comparison to women with normal pre-pregnancy BMI by 34.9% (AOR=0.651, CI=0.482-0.878, p=0.005). Presence of co-morbidities among the COVID-19 positive pregnant women increased the chance of undergoing cesarean section by 53.4% (AOR=1.534, CI=1.210-1.945, p<0.001) (Table 4).

**Table 4:**
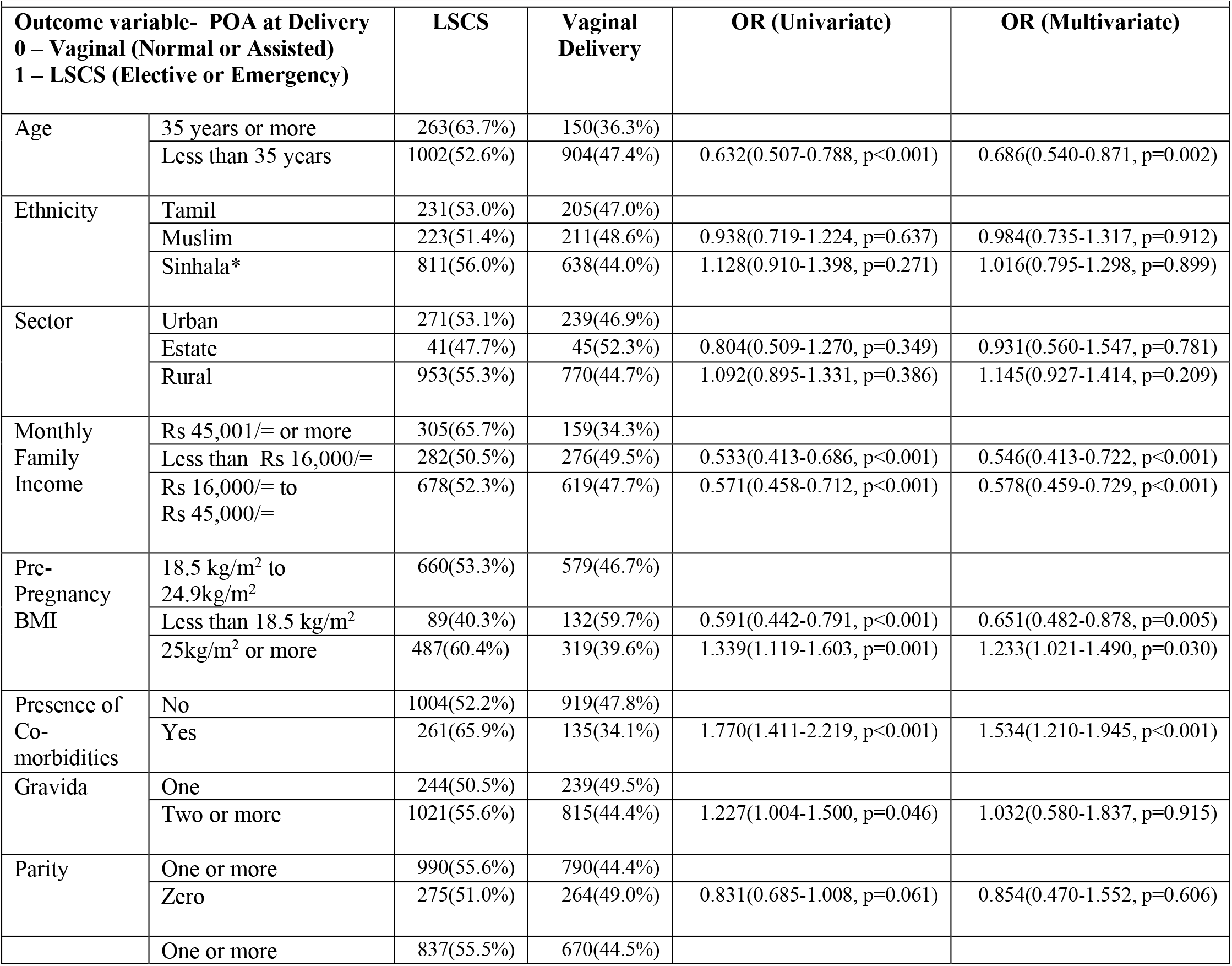
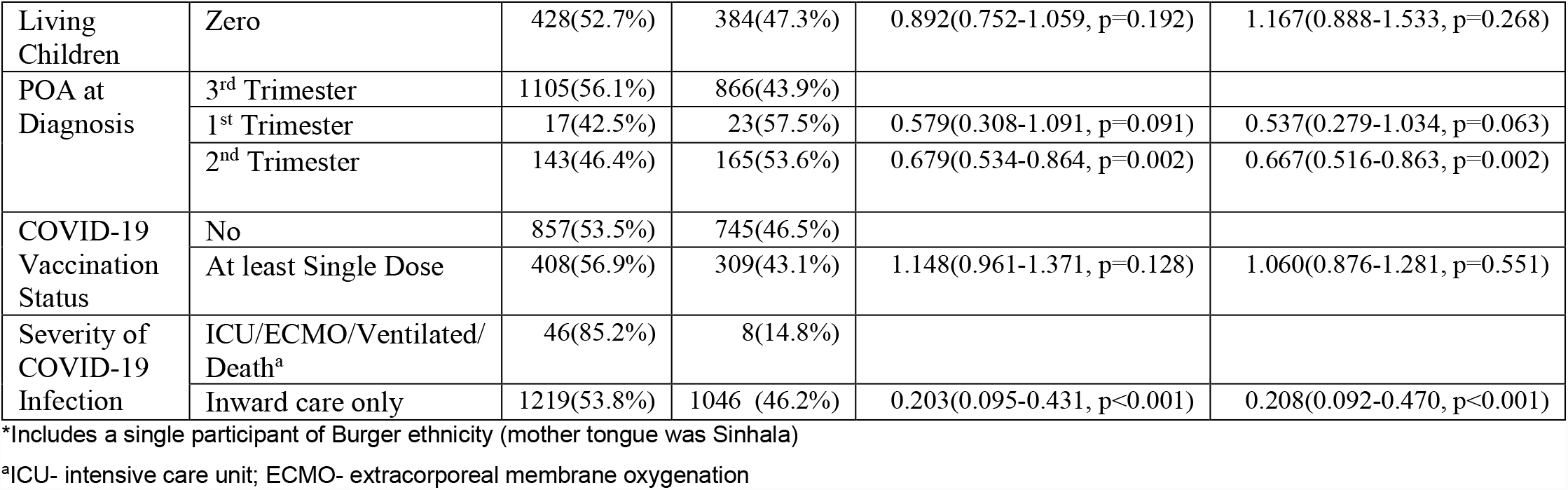
Odds Ratios for factors associated with mode of delivery of COVID-19 positive pregnant women. Univariate and multivariate analysis.

### Factors associated with birthweight of the newborns of COVID-19 positive pregnant women

Compared to COVID-19 positive pregnant women of 35 years or older, pregnant women aged less than 35 years were less likely to give birth to newborns with low birthweight by 44.6% (AOR=0.554, CI=0.415-0.738, p<0.001). In contrast, COVID-19 positive pregnant women belonging to estate sector displayed increased risk of giving birth to low birthweight newborns compared to unban pregnant women (AOR=2.413, CI=1.344-4.334, p=0.003). Similarly, COVID-19 positive women with low pre-pregnancy BMI demonstrated increased risk compared to women with normal pre-pregnancy BMI by 74.6% (AOR=1.746, CI=1.231-2.476, p=0.002). In contrast the risk of low birthweight reduced by 69.9% among the COVID-19 positive women who required only inward treatment only compared to the women needing ICU care or death due to COVID-19 (AOR=0.301, CI=0.165-0.551, p<0.001) (Table 5).

**Table 5.** Odds Ratios for factors associated with birthweight of newborns of COVID-19 positive pregnant women. Univariate and multivariate analysis.

|  |  |  |  |  |  |
| --- | --- | --- | --- | --- | --- |
| **Outcome variable- Birthweight** |  | **2500g or more** | **Less than 2500g** | **OR (Univariate)** | **OR (Multivariate)** |
| **0 – 2500g or more** |  |  |  |  |  |
| **1 – Less than 2500g** |  |  |  |  |  |
| Age | 35 years or more | 317(77.1%) | 94(22.9%) |  |  |
|  | Less than 35 years | 1617(84.8%) | 289(15.2%) | 0.603(0.464-0.783, p<0.001) | 0.554(0.415-0.738, p<0.001) |
| Ethnicity | Tamil | 345(79.1%) | 91(20.9%) |  |  |
|  | Muslim | 366(84.7%) | 66(15.3%) | 0.684(0.482-0.970, p=0.033) | 0.917(0.626-1.345, p=0.659) |
|  | Sinhala* | 1223(84.4%) | 226(15.6%) | 0.701(0.534-0.919, p=0.010) | 0.827(0.603-1.133, p=0.237) |
| Sector | Urban | 442(86.7%) | 68(13.3%) |  |  |
|  | Estate | 60(69.8%) | 26(30.2%) | 2.817(1.664-4.767, p<0.001) | 2.413(1.344-4.334, p=0.003) |
|  | Rural | 1432(83.2%) | 289(16.8%) | 1.312(0.987-1.744, p=0.062) | 1.277(0.949-1.718, p=0.107) |
| Monthly Family Income | Rs 45,001/= or more | 392(84.7%) | 71(15.3%) |  |  |
|  | Less than Rs 16,000/= | 449(80.5%) | 109(19.5%) | 1.340(0.965-1.861, p=0.080) | 1.176(0.817-1.693, p=0.382) |
|  | Rs 16,000/= to Rs 45,000/= | 1093(84.3%) | 203(15.7%) | 1.025(0.764-1.376, p=0.867) | 1.042(0.764-1.422, p=0.793) |
| Pre-Pregnancy BMI | 18.5 kg/m<sup>2</sup> to 24.9kg/m<sup>2</sup> | 1043(84.2%) | 196(15.8%) |  |  |
|  | Less than 18.5 kg/m<sup>2</sup> | 165(74.7%) | 56(25.3%) | 1.806(1.286-2.536, p=0.001) | 1.746(1.231-2.476, p=0.002) |
|  | 25kg/m<sup>2</sup> or more | 683(85.0%) | 121(15.0%) | 0.943(0.737-1.206, p=0.639) | 0.912(0.705-1.179, p=0.482) |
| Presence of Co-morbidities | No | 1608(83.7%) | 313(16.3%) |  |  |
|  | Yes | 326(82.3%) | 70(17.7%) | 1.103(0.829-1.467, p=0.500) | 1.093(0.806-1.482, p=0.565) |
| Gravida | One | 397(82.2%) | 86(17.8%) |  |  |
|  | Two or more | 1537(83.8%) | 297(16.2%) | 0.892(0.685-1.162, p=0.397) | 0.868(0.405-1.861, p=0.717) |
| Parity | One or more | 1490(83.8%) | 288(16.2%) |  |  |
|  | Zero | 444(82.4%) | 95(17.6%) | 1.107(0.858-1.428, p=0.435) | 0.925(0.420-2.036, p=0.847) |
| Living Children | One or more | 1265(84.1%) | 240(15.9%) |  |  |
|  | Zero | 669(82.4%) | 143(17.6%) | 1.127(0.898-1.414, p=0.304) | 1.240(0.867-1.774, p=0.240) |
| POA at Diagnosis | 3<sup>rd</sup> Trimester | 1643(83.4%) | 326(16.6%) |  |  |
|  | 1<sup>st</sup> Trimester | 34(85.0%) | 6(15.0%) | 0.889(0.370-2.136, p=0.793) | 0.970(0.397-2.372, p=0.947) |
|  | 2<sup>nd</sup> Trimester | 257(83.4%) | 51(16.6%) | 1.000(0.724-1.382, p=0.999) | 1.107(0.785-1.562, p=0.562) |
| COVID-19 Vaccination Status | No | 1643(83.4%) | 326(16.6%) |  |  |
|  | At least Single Dose | 588(82.2%) | 127(17.8%) | 1.136(0.899-1.435, p=0.286) | 1.113(0.866-1.429, p=0.403) |
| Severity of COVID-19 Infection | ICU/ECMO/Ventilated/Death<sup>a</sup> | 34(63.0%) | 20(37.0%) |  |  |
|  | Inward care only | 1900(84.0%) | 363(16.0%) | 0.325(0.185-0.571, p<0.001) | 0.301(0.165-0.551, p<0.001) |

\*Includes a single participant of Burger ethnicity (mother tongue was Sinhala)
<sup>a</sup>ICU- intensive care unit; ECMO- extracorporeal membrane oxygenation

### Factors associated with presence of congenital abnormalities of newborns of COVID-19 positive pregnant women

Among the COVID-19 positive women who required only inward treatment, presence of any type of congenital abnormalities in newborn reduced by 72.4% in comparison to women with severe COVID-19 infection (AOR=0.276, CI=0.112-0.683, p=0.005) (Table 6).

**Table 6.** Odds Ratios for factors associated with the presence of congenital abnormalities of neonate of COVID-19 positive pregnant women. Univariate and multivariate analysis.

| Outcome variable- Congenital abnormalities<br>0 - Absent<br>1 – Present |  | Absent | Present | OR (Univariate) | OR (Multivariate) |
| --- | --- | --- | --- | --- | --- |
| Age | 35 years or more | 399(95.5%) | 19(4.5%) |  |  |
|  | Less than 35 years | 1866(97.1%) | 56(2.9%) | 0.630(0.370-1.072, p=0.089) | 0.656(0.371-1.161, p=0.148) |
| Ethnicity | Tamil | 431(97.5%) | 11(2.5%) |  |  |
|  | Muslim | 425(96.8%) | 14(3.2%) | 1.291(0.579-2.875, p=0.532) | 1.399(0.604-3.242, p=0.433) |
|  | Sinhala* | 1409(96.6%) | 50(3.4%) | 1.390(0.717-2.694, p=0.329) | 1.341(0.649-2.771, p=0.429) |
| Monthly Family Income | Rs 45,001/= or more | 455(97.2%) | 13(2.8%) |  |  |
|  | Less than Rs 16,000/= | 547(97.2%) | 16(2.8%) | 1.024(0.487-2.151, p=0.951) | 1.296(0.582-2.886, p=0.526) |
|  | Rs 16,000/= to Rs 45,000/= | 1263(96.5%) | 46(3.5%) | 1.275(0.682-2.381, p=0.446) | 1.436(0.742-2.780, p=0.282) |
| Pre-Pregnancy BMI | 18.5 kg/m<sup>2</sup> to 24.9kg/m<sup>2</sup> | 1209(96.9%) | 39(3.1%) |  |  |
|  | Less than 18.5 kg/m<sup>2</sup> | 216(97.3%) | 6(2.7%) | 0.861(0.360-2.059, p=0.737) | 0.982(0.405-2.383, p=0.969) |
|  | 25kg/m<sup>2</sup> or more | 788(96.7%) | 27(3.3%) | 1.062(0.645-1.749, p=0.813) | 0.902(0.536-1.519, p=0.698) |
| Presence of Co-morbidities | No | 1883(97.3%) | 53(2.7%) |  |  |
|  | Yes | 382(94.6%) | 22(5.4%) | 2.046(1.230-3.404, p=0.006) | 1.813(1.046-3.143, p=0.034) |
| Gravida | One | 473(97.5%) | 12(2.5%) |  |  |
|  | Two or more | 1792(96.6%) | 63(3.4%) | 1.386(0.741-2.590, p=0.307) | 0.662(0.082-5.324, p=0.698) |
| Parity | One or more | 1736(96.6%) | 62(3.4%) |  |  |
|  | Zero | 529(97.6%) | 13(2.4%) | 0.688(0.375-1.261, p=0.226) | 0.528(0.064-4.373, p=0.554) |
| Living Children | One or more | 1467(96.5%) | 53(3.5%) |  |  |
|  | Zero | 798(97.3%) | 22(2.7%) | 0.763(0.461-1.294, p=0.294) | 0.936(0.429-2.042, p=0.868) |
| POA at Diagnosis | 3<sup>rd</sup> Trimester | 1929(97.2%) | 56(2.8%) |  |  |
|  | 1<sup>st</sup> Trimester | 37(92.5%) | 3(7.5%) | 2.793(0.836-9.331, p=0.095) | 2.849(0.835-9.722, p=0.095) |
|  | 2<sup>nd</sup> Trimester | 299(94.9%) | 16(5.1%) | 1.843(1.044-3.255, p=0.035) | 1.811(0.978-3.352, p=0.059) |
| COVID-19 Vaccination Status | No | 1561(96.7%) | 54(3.3%) |  |  |
|  | At least Single Dose | 704(97.1%) | 21(2.9%) | 0.862(0.517-1.439, p=0.570) | 0.962(0.555-1.667, p=0.890) |
| Severity of COVID-19 Infection | ICU/ECMO/Ventilated/Death<sup>a</sup> | 53(86.9%) | 8(13.1%) |  |  |
|  | Inward care only | 2212(97.1%) | 67(2.9%) | 0.201(0.092-0.439, p<0.001) | 0.276(0.112-0.683, p=0.005) |

\*Includes a single participant of Burger ethnicity (mother tongue was Sinhala)
<sup>a</sup>ICU- intensive care unit; ECMO- extracorporeal membrane oxygenation

### Factors associated with the place of immediate postnatal management of the newborn of COVID-19 positive pregnant women

Table seven demonstrates factors associated with the place of immediate management of the newborn of COVID-19 positive pregnant women of Sri Lanka. High pre-pregnancy BMI increased the risk of newborn admitting to PBU or NICU in comparison to pregnant women with normal BMI by 34.9% (AOR=0.651, CI=0.466-0.909, p=0.012). In contrast, among the COVID-19 positive pregnant women only requiring inward care, the chances of the newborns only managed at postnatal ward increased significantly (AOR=11.575, CI=6.480-20.678, p<0.001) (Table 7).

**Table 7.** Odds Ratios for factors associated with the place of immediate postnatal management of the newborn of COVID-19 positive pregnant women. Univariate and multivariate analysis.

|  |  |  |  |  |  |
| --- | --- | --- | --- | --- | --- |
| **Outcome variable- Place of management of the newborn**<br>**0 – Premature Baby Unit/ Neonatal ICU (PBU/ NICU)**<br>**1 – Postnatal ward only** |  | **PBU/ NICU** | **Postnatal ward only** | **OR (Univariate)** | **OR (Multivariate)** |
| Age | 35 years or more | 43(10.4%) | 372(89.6%) |  |  |
|  | Less than 35 years | 151(7.9%) | 1763(92.1%) | 1.350(0.945-1.928, p=0.100) | 1.357(0.909-2.028, p=0.136) |
| Ethnicity | Tamil | 38(8.7%) | 401(91.3%) |  |  |
|  | Muslim | 26(5.9%) | 411(94.1%) | 1.498(0.893-2.513, p=0.126) | 1.418(0.806-2.496, p=0.225) |
|  | Sinhala* | 130(8.9%) | 1323(91.1%) | 0.964(0.661-1.408, p=0.851) | 0.951(0.613-1.476, p=0.824) |
| Sector | Urban | 51(10.0%) | 461(90.0%) |  |  |
|  | Estate | 7(8.1%) | 79(91.9%) | 1.249(0.547-2.850, p=0.598) | 1.262(0.509-3.129, p=0.615) |
|  | Rural | 136(7.9%) | 1595(92.1%) | 1.297(0.925-1.819, p=0.131) | 1.358(0.946-1.949, p=0.098) |
| Monthly Family Income | Rs 45,001/= or more | 43(9.2%) | 422(90.8%) |  |  |
|  | Less than Rs 16,000/= | 49(8.7%) | 513(91.3%) | 1.067(0.694-1.639, p=0.768) | 0.802(0.493-1.305, p=0.375) |
|  | Rs 16,000/= to Rs 45,000/= | 102(7.8%) | 1200(92.2%) | 1.199(0.825-1.741, p=0.341) | 1.020(0.680-1.531, p=0.923) |
| Pre-Pregnancy BMI | 18.5 kg/m<sup>2</sup> to 24.9kg/m<sup>2</sup> | 88(7.1%) | 1155(92.9%) |  |  |
|  | Less than 18.5 kg/m<sup>2</sup> | 15(6.8%) | 207(93.2%) | 1.051(0.596-1.854, p=0.862) | 1.034(0.577-1.852, p=0.911) |
|  | 25kg/m<sup>2</sup> or more | 85(10.7%) | 725(89.5%) | 0.650(0.476-0.888, p=0.007) | 0.651(0.466-0.909, p=0.012) |
| Presence of Co-morbidities | No | 153(7.9%) | 1776(92.1%) |  |  |
|  | Yes | 41(10.3%) | 359(89.8%) | 0.754(0.525-1.084, p=0.128) | 0.929(0.624-1.383, p=0.717) |
| Gravida | One | 41(8.5%) | 444(91.5%) |  |  |
|  | Two or more | 153(8.3%) | 1691(91.7%) | 1.021(0.712-1.463, p=0.912) | 0.683(0.289-1.619, p=0.387) |
| Parity | One or more | 145(8.1%) | 1642(91.9%) |  |  |
|  | Zero | 49(9.0%) | 493(91.0%) | 0.888(0.633-1.247, p=0.494) | 0.785(0.321-1.916, p=0.594) |
| Living Children | One or more | 116(7.7%) | 1397(92.3%) |  |  |
|  | Zero | 78(9.6%) | 738(90.4%) | 0.786(0.582-1.061, p=0.116) | 0.621(0.390-0.989, p=0.045) |
| POA at Diagnosis | 3<sup>rd</sup> Trimester | 165(8.3%) | 1812(91.7%) |  |  |
|  | 1<sup>st</sup> Trimester | 3(7.5%) | 37(92.5%) | 1.123(0.343-3.682, p=0.848) | 1.115(0.323-3.854, p=0.863) |
|  | 2<sup>nd</sup> Trimester | 26(8.3%) | 286(91.7%) | 1.002(0.650-1.543, p=0.994) | 0.980(0.614-1.564, p=0.932) |
| COVID-19 Vaccination Status | No | 136(8.5%) | 1471(91.5%) |  |  |
|  | At least Single Dose | 58(8.0%) | 664(92.0%) | 1.058(0.768-1.459, p=0.729) | 1.043(0.738-1.475, p=0.812) |
| Severity of COVID-19 Infection | ICU/ECMO/Ventilated/Death<sup>a</sup> | 28(50.0%) | 28(50.0%) |  |  |
|  | Inward care only | 166(7.3%) | 2107(92.7%) | 12.693(7.344-21.937, p<0.001) | 11.575(6.480-20.678, p<0.001) |

\*Includes a single participant of Burger ethnicity (mother tongue was Sinhala)
<sup>a</sup>ICU- intensive care unit; ECMO- extracorporeal membrane oxygenation

## DISCUSSION

### Principal findings

Analysis of pregnancy and neonatal characteristics of COVID-19 positive pregnant women of Sri Lanka revealed pre-pregnancy overweight rate of 35.7% with only 31% receiving at least single dose of any type of COVID-19 vaccine. Presence of gestational DM was at 8% and pregnancy induced hypertension at 3% among the COVID-19 positive pregnant women. Maternal COVID-19 infection reported preterm birth rate of 11.9% and LSCS rate of 54.5%. Low birthweight rate was 16.5% with 8.3% of the newborns requiring intensive care among these women. Neonatal mortality rate was reported at 9 per 1000 live births among the COVID-19 positive pregnant women. Pre-pregnancy overweight and obesity increased the risk of preterm delivery among the COVID-19 positive women and increased the probability of undergoing cesarean section compared to normal BMI women. Similarly, presence of co-morbidities among these women increased the probability of undergoing cesarean section compared to women with no co-morbidities. COVID-19 positive pregnant women belonging to estate sector displayed increased risk of giving birth to low birthweight newborns compared to unban pregnant women. The risk of low birthweight reduced by 69.9% among the COVID-19 positive women who required only inward treatment only compared to the women needing ICU care or death due to COVID-19. In addition, high pre-pregnancy BMI increased the risk of newborn admitting to PBU or NICU in comparison to pregnant women with normal BMI by 34.9%.

### National COVID-19 positive pregnant women surveillance

The national COVID-19 positive pregnant women surveillance in Sri Lanka reported 9,441 cases of maternal COVID-19 was reported of which only 157 were reported in 2020. Majority of the maternal COVID-19 cases were reported from August to October of 2021 (6,662).[15] Therefore, delivery outcome of majority of these pregnant women were not reported by 31st of October which resulted in inclusion of only 2340 maternal COVID-19 cases out of 9,441. Number of maternal COVID-19 cases who were tested positive and not reported in the national surveillance is negligible due to several reasons. Centralized reporting of COVID-19 positive cases from all laboratories (both private and government) in Sri Lanka and the surveillance mechanism which forward these information to the relevant Medical Officer of Health areas for patient tracing and quarantine measures ensure that the field public health workers are updated on all COVID-19 positive cases including pregnant women. In addition, close to 95% of pregnant women of Sri Lanka have received domiciliary care by PHMs in 2019 which would have further reduced the possibility of missing positive maternal cases.[15]

### Socio-demographic and maternal characteristics

As reported by Kumarasinghe et al., percentage of pregnant women who were 35 years or older was 13.6% in Sri Lanka in 2020-2021 whereas present study revealed 17.9% of pregnant women in same age group highlighting the increase incidence of symptomatic COVID-19 infection with advancing age in Sri Lanka.[15] However, studies in America and Italy have suggested significant decrease in incidence of COVID-19 among the 35 years or above category in similar population based studies.[16-17] Onset and coverage of vaccination of pregnant women could be a factor in the observed difference. Though the countries like America and Italy initialed vaccinating pregnant women earlier than Sri Lanka, the coverage is an issue with large number of youth avoiding vaccination in America and Europe.[18-19]. In Sri Lanka, it is reported that around 80% to 90% of pregnant women in Sri Lanka was vaccinated against COVID-19 by September of 2021 being the first country in South Asia to commence vaccinating pregnant women (Among the COVID-19 positive women only 31% have received at least a single dose of any recommended vaccine before 14 days of the diagnosis.[20]

### Preterm delivery

Preterm delivery rate(less than 37 weeks) in the present study among COVID-19 positive pregnant women was 11.9%. Published national preterm rates are not available for Sri Lanka. Many studies reported higher rates of preterm delivery among COVID-19 positive women compared to the present study. However, these studies were not population based studies and were confined to a single or multiple centers. These studies have only included pregnant women who were either tested positive on admission for the delivery or delivered during the hospital stay following admission to hospital with the diagnosis of COVID-19. Pre-term delivery rates ranged from 13.8% to 59.4%.[8, 21-25]

Present study showed increased risk of preterm delivery among the pregnant women with severe COVID-19 infection. Conflicting evidence have emerged on whether the severity of the maternal COVID-19 infection influence the preterm delivery. A retrospective multicenter study in United States concluded that acute COVID-19 severity was not correlated with gestational age at delivery whereas separate cohort study in United States reported significant increase in preterm delivery among critical group compared to severe group.[22, 26] Our study reported preterm delivery rate of 40.7% among pregnant women needing ICU/ ECMO/ ventilated/ death whereas Pierce-Williams reported preterm delivery rate of 27% among severe group and 88% among critical group.[22]

### Mode of Delivery

Preferred mode of delivery of the COVID-19 positive pregnant women in Sri Lanka was cesarean section with 54.5% undergoing CS. The cesarean section rate of this study is among the one the highest among the published studies with large sample sizes. Not only among COVID-19 positive pregnant women, but national CS rate in Sri Lanka is also one of the highest in the world which might have contributed the increased CS rate of maternal COVID-19.[27-28] The CS rate among COVID-19 positive pregnant women ranged from 22.4% in America to 94% in China. Some of these studies included less than 100 respondents.[8, 21-23, 26]

Similar to the general population of pregnant women, pre-pregnancy overweight and obesity increased the chances of cesarean section among the maternal COVID-19 positive women in our study population. Significant increase in the CS rates were observed in United States cohort study with increase in severity of maternal COVID-19 infection in consistent with our study findings.[22]

### Birthweight of the newborn

Present study revealed low birthweight of 16.5%. Range of low birthweight reported by studies ranged from 5% to 67%. Many reasons could have contributed to this wide range. One could be the characteristics of the sample including the proportion of severe to critical women. Second, the inclusion criteria; Whether all women of past and present COVID-19 infection or only the women with active infection during the delivery was included. Third, the study setting, with some were institutional based whereas few were population based. Sample size also could have played a role in difference. However, it is important to compare the reported low birthweight rates with the national averages as many factors including sociodemographic factors play an important role in determining the low birthweight among COVID-19 positive pregnant women. Sri Lanka reported low birthweight of 16% in 2018.[28] Thus, only 0.5% increase in low birthweight was observed in the present study.[7-8, 25, 29-32]

Increase severity was associated with higher low birthweight rates as concluded by studies including the present one.[22] Our study revealed significantly higher rates of low birth weight among older pregnant women which is a known risk factor for low birthweight irrespective of the presence of COVID-19 among pregnant women. However, Dubey et al. in their meta-analysis failed to demonstrate statistical difference though the age categorization differed from the present study (35 or more vs. more than 30 years).[8, 33]

### Immediate Postnatal care and presence of congenital abnormalities

As described under “birthweight of the newborn” many variations in study design, population, study setting and sample size have contributed to wide range of reporting rates of intensive postnatal care among maternal COVID-19 infection from 8.3% (present study) to 76.9%.[22-23, 30, 32, 34-35]

Similar to present study, increased severity was associated with higher possibility of admission to intensive care for postnatal management of the newborn.[22] Further present study reported newborns of overweight/ obese pregnant women are at higher probability of admission to intensive care postnatally. Pre-pregnancy obesity in a known independent risk factor for neonatal intensive care admission. Therefore, the observed increased neonatal admission could be due to overweight/ obesity irrespective of the maternal COVID-19 status.[36]

Our study reported incidence of congenital abnormalities of any type and severity at 3.2%. The national birth effect rate was less than 2%. Therefore, there could be a possibility of increased incidence of congenital abnormalities among the newborns of COVID-19 positive pregnant women. At present, limited literature is available that provide clear direction on congenital abnormalities and COVID. Thus, further research is needed to have a clear picture.[37-38]

## CONCLUSION

Increased severity of COVID-19 pregnant women was associated with many adverse pregnancy and neonatal outcomes including increased cesarean section rates, preterm delivery, low birthweight and congenital abnormalities compared to less severe infection. Similarly, pre-pregnancy overweight/ obesity could increase the risk of many negative pregnancy and neonatal outcomes compared to pregnant women with normal BMI. Thus, close observation and management of COVID-19 among the pregnant women should be considered to reduce the risk of progressing to severe illness. This could have an impact in reducing the adverse pregnancy and neonatal outcomes among maternal COVID-infection.

## Data Availability

All data produced in the present study are available upon reasonable request to the authors

## Acknowledgements

We acknowledge all the front-line public health staff, namely field Public Health Midwives, Public Health Nursing Sisters, Medical Officers of Health and regional health managers, namely Medical Officers-Maternal and Child Health for their contribution for the National COVID-19 Positive Pregnant Women Surveillance.

## Contributors

HJ and KK developed the study protocol and overall workplan with JD and SG. MK was responsible for data extraction, cleaning, and analysis with the assistance of SI and WS. MK led the writing with all the authors contributed for writing and reviewing the final manuscript. CdS supervised the overall the project.

## Disclaimer

The views expressed are those of the authors and not necessarily those of the Family Health Bureau, Sri Lanka or Ministry of Health, Sri Lanka

## Patient consent for publication

Not applicable

## Ethics approval

Ethical approval was obtained from Ethics Review Committee of Faculty of Medical Sciences, University of Sri Jayawardenepura (COVID 11/21).

## Data availability statement

Data are available on reasonable request. Raw data without personal identifiers are available from corresponding author on reasonable request.

